# Trends in Industry-Sponsored Research Funding to Orthopaedic Surgeons in the U.S. from 2017-2024

**DOI:** 10.1101/2025.11.07.25339785

**Authors:** Justin S. King, Abdullah H. Salama, Archana Sanjay, Michael H. Weber, Isaac L. Moss

## Abstract

Orthopaedic injuries and musculoskeletal (MSK) diseases limit function, reduce quality of life, and carry substantial socioeconomic burdens. Because federal support for orthopaedic research is disproportionately low and volatile, industry-sponsored research funding (ISRF) can provide additional support to advance clinical care. We performed an analysis of ISRF directed to orthopaedic surgeons Principal Investigators (PIs) between 2017 and 2024 using the Centers for Medicare & Medicaid Services Open Payments database. Payments were aggregated per PI, then stratified by subspecialty, study phase (pre-clinical vs registered clinical trial), and sponsoring company. In doing so, we found that orthopaedic surgeon PIs received $395.7 million in ISRF during this eight-year span; each year this averaged $49.5 million from 106 companies to 956 investigators, with the median ISRF $14,121 to each PI. Across the study period, approximately 5% of ISRF supported pre-clinical work, and 20% was tied to registered clinical trials. Surgeons who received ISRF also accepted higher general (non-research) industry payments than their non-research peers ($69,441 vs $8,363; p<0.001). Together, these results show that ISRF provides an avenue to conduct research and augment other sources of funding.

**Clinical Significance:** Understanding ISRF patterns can help academic centers and professional societies anticipate future funding directions, direct resources, and help orthopaedic surgeons acquire funding that advances care and benefits patients.

## Introduction

Musculoskeletal (MSK) diseases, such as arthritis, back pain, ligamentous injuries, and fractures, affect approximately 41% of the United States (US) population. The resulting morbidity and mortality leads to direct healthcare expenditures equating to about $380.9 billion annually, not including indirect costs (1). When assessed by disability-adjusted life years (DALYs), MSK conditions are the leading cause of disability and disease burden in the US (2–4). Despite the socioeconomic impact of orthopaedic conditions, federal research funding is disproportionately low. A recent analysis showed that federal research funding for MSK-related research was about $351 million per year, compared to cancer receiving $6.5 billion per year. As the population ages, the burden of MSK conditions will rise, representing an urgent need for more effective, affordable therapies that will depend on funded research. This research may be funded by government, non-profit, and industry sources (5–9).

To explore the dynamics of industry-sponsored research funding (ISRF) directed to orthopaedic surgeons, we utilized data from the Centers for Medicaid Services (CMS) Open Payments database (OBD), which requires the disclosure of financial transactions between industry and practicing physicians as mandated by the Physician Payments Sunshine Act. These disclosures largely apply to organizations that colloquially include biotech, device, and pharmaceutical companies. These disclosures are updated annually and released by June 30th for the previous year. This reporting requirement aims to promote transparency and highlight the potential impact of financial relationships between healthcare providers and industry partners (10).

OPD disclosures are broadly categorized as *General Payments*, *Ownership or Investment Interest*s, and *Research Payments* (ISRF) (11). *General Payments* include most non-research-related items such as consulting fees, food, royalties, and speaking fees*. Ownership or Investment Interests* refer to financial interests held by physicians in the reporting companies (12). ISRF is typically directed to institutions, such as teaching hospitals, as opposed to individuals, but if the principal investigators (PIs) who are leading the research are a healthcare provider, these payments must also be disclosed in the Open Payments database (13).

Orthopaedic surgeons have consistently been among the highest recipients of industry funding within the *General Payments* category (8,10); however, fewer studies have assessed ISRF, with the most recent analysis through the 2021 reporting year showing that orthopaedic-surgeon principal investigators received about $46.8 million in ISRF (8,9). While considerable, total ISRF reported through OPD in 2021 was $7.82 billion, which has grown to $8.52 billion in 2024 (8,11). However, it remains unclear how the distribution and trends of ISRF to orthopaedic surgeons have evolved in recent years, especially amid overall national growth in industry funding. Therefore, this study aims to characterize temporal trends, funding magnitude, and subspecialty distribution of ISRF to orthopaedic surgeons between 2017 and 2024. We hypothesize that industry-sponsored research funding to orthopaedic surgeons has increased over this period but remains disproportionately low relative to the burden of disease. We also wanted to assess if ISRF was concentrated among specific subspecialties, so that orthopaedic surgeons can better position themselves to secure research support.

## Methods

### Data Source

This study utilizes data from the Open Payments database, maintained by the Centers for Medicaid Services (CMS) (11). We extracted data related to Research and General Payments made to those with a designated primary specialty as *orthopaedic surgeon* from 2017 to 2024. For Research Payments, the orthopaedic surgeon was listed as the Primary Principal Investigator (PI) and all payments under this category were noted as industry-sponsored research funding (ISRF). For General Payments, they were included as the Primary Receipt.

### Data Extraction and Preparation

To consolidate entries where multiple payments were made to the same individual, we matched them based on National Provider Identifier (NPI) numbers. Orthopaedic surgeons were further classified into various subspecialties based on the designated specialty in the CMS dataset. For reporting industry-related funding, we aggregated payments by unique healthcare organization or Group Purchasing Organization (GPO) identifier, collectively referred to as industry sponsors. Dollar values were rounded to the nearest whole number. We matched General and Research payments based on NPI. Race/ethnicity information is not included in disclosures.

### Statistical Analysis

Annual totals and medians were reported for each subspecialty from 2017 through 2024. Inter-quartile range (IQR) for first and third quartile are report where indicated. Because payment distributions were highly skewed, non-parametric tests were used. Kruskal– Wallis test indicated overall group differences, all pairwise Wilcoxon rank-sum tests were conducted, and p-values were adjusted for multiple comparisons using the Bonferroni method. Data processing and analysis were performed using R (4.3.1; Posit, PBC).

## Results

### Research payment trends

From 2017 to 2024, industry-sponsored research funding (ISRF) directed to orthopaedic surgeon principal investigators (PIs) totaled $395.7 million, averaging $49.5 million annually (Figure 1A). Each year, funding was distributed to an average of 956 PIs, with a median ISRF of $14,121 (IQR: $4,285–$41,734) per investigator. However, the distribution of ISRF was highly skewed, with the top 5 % of orthopaedic surgeon–researchers receiving 52 % of total funding, amounting to at least $170,665 per year (Figure 1B).

**Figure 1.**
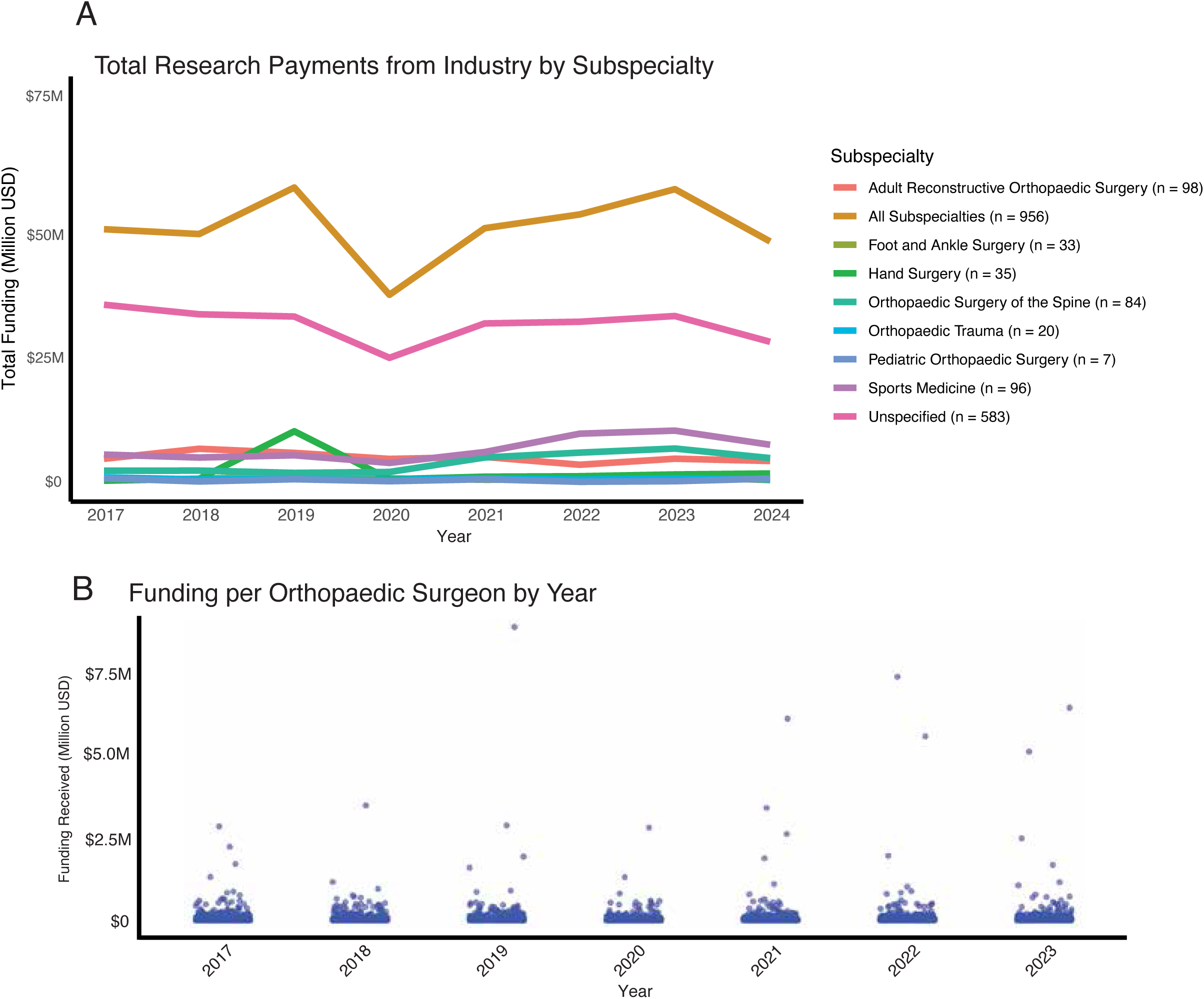
Trends in industry payments to orthopaedic surgeons. (A) Total research funding made to orthopaedic surgeons broken down by listed subspeciality with the average number (n) of orthopaedic surgeons in each specialty. (B) Each dot represents an individual orthopaedic surgeon and the amount of research funding they received in each year listed.

Across subspecialties, the largest total ISRF was directed to investigators without a listed subspecialty (“unspecified,” n = 583), followed by those in Sports Medicine, Adult Reconstructive Surgery, and Spine Surgery. This difference was largely due to a higher number of funded investigators in these subspecialities as the per investigator, median ISRF levels was generally similar across subspecialties. Notable exceptions included Pediatric Orthopaedic Surgery ($29,520; IQR $12,612–$54,475; n = 7), which, despite only seven funded investigators, had the highest per-PI support, and Foot & Ankle Surgery ($9,340; IQR $4,069–$20,917; n = 33), which had the lowest. Other subspecialties showed intermediate levels, including Sports Medicine ($17,174; IQR $5,250–$51,102; n = 96), Adult Reconstructive Surgery ($16,627; IQR $4,854–$50,458; n = 98), Orthopaedic Trauma ($14,983; IQR $6,001–$33,870; n = 20), Spine Surgery ($14,739; IQR $4,671–$40,848; n = 84), Unspecified ($13,990; IQR $4,277–$41,251; n = 583), and Hand Surgery ($11,970; IQR $4,227–$34,052; n = 35) (Table 1).

**Table 1:** Payment to orthopaedic surgeons broken down by subspecialty.

| Subspecialty / Year | 2017 | 2018 | 2019 | 2020 | 2021 | 2022 | 2023 | 2024 | Average |
| --- | --- | --- | --- | --- | --- | --- | --- | --- | --- |
| Pediatrics | \$15,300 | \$9,700 | \$81,550 | \$19,215 | \$16,083 | \$19,592 | \$17,886 | \$56,835 | \$29,520 |
| Sports Medicine | \$13,469 | \$17,274 | \$16,003 | \$15,601 | \$14,506 | \$21,850 | \$18,694 | \$20,000 | \$17,174 |
| Adult Reconstructive | \$19,694 | \$18,909 | \$23,273 | \$12,035 | \$15,300 | \$14,259 | \$16,236 | \$13,308 | \$16,627 |
| Orthopaedic Trauma | \$30,014 | \$17,187 | \$11,864 | \$11,000 | \$4,348 | \$18,000 | \$13,301 | \$14,148 | \$14,983 |
| Spine | \$11,784 | \$14,350 | \$10,151 | \$12,500 | \$9,728 | \$17,589 | \$23,210 | \$18,600 | \$14,739 |
| Unspecified | \$14,050 | \$16,728 | \$15,063 | \$11,844 | \$11,496 | \$12,933 | \$14,737 | \$15,070 | \$13,990 |
| Hand Surgery | \$8,528 | \$11,049 | \$9,552 | \$4,612 | \$9,125 | \$10,307 | \$20,883 | \$21,703 | \$11,970 |
| Foot and Ankle | \$12,133 | \$10,350 | \$10,750 | \$11,755 | \$6,670 | \$8,881 | \$6,185 | \$7,995 | \$9,340 |
| *Funding reported in United States Dollars |  |  |  |  |  |  |  |  |  |

A Kruskal–Wallis test revealed significant differences in median ISRF per PI across orthopaedic subspecialties in 2024 (p = 0.0027). Post-hoc Wilcoxon rank-sum tests with Bonferroni correction identified that Hand Surgery differed significantly from Foot & Ankle Surgery (p = 0.020), Spine Surgery (p = 0.020), and Pediatric Orthopaedic Surgery (p = 0.010). Foot & Ankle Surgery also differed significantly from Sports Medicine (p = 0.045) and Pediatric Orthopaedics (p = 0.010). No other pairwise comparisons reached statistical significance (p > 0.05), indicating that although total funding varied by subspecialty, per-investigator differences were generally similar.

### Uses of the payments received

Open Payments records indicate whether a payment supports pre-clinical research or a registered clinical trial. The annual proportion of funding attributed to clinical trials ranged from a low of 13% in 2022 and 2023 to a peak of 32% in 2019. In 2019 alone, payments related to clinical trials totaled $18.3 million across 112 unique trials and 321 individual investigators. However, this dropped in 2020 to 17% ($6.3 million). Funding explicitly labelled “pre-clinical research” typically accounted for 1.9-4.4 % of annual ISRF and reached its highest level in 2023, when $2.5 million (4.4%) was distributed to 63 investigators (Table 2).

**Table 2.**
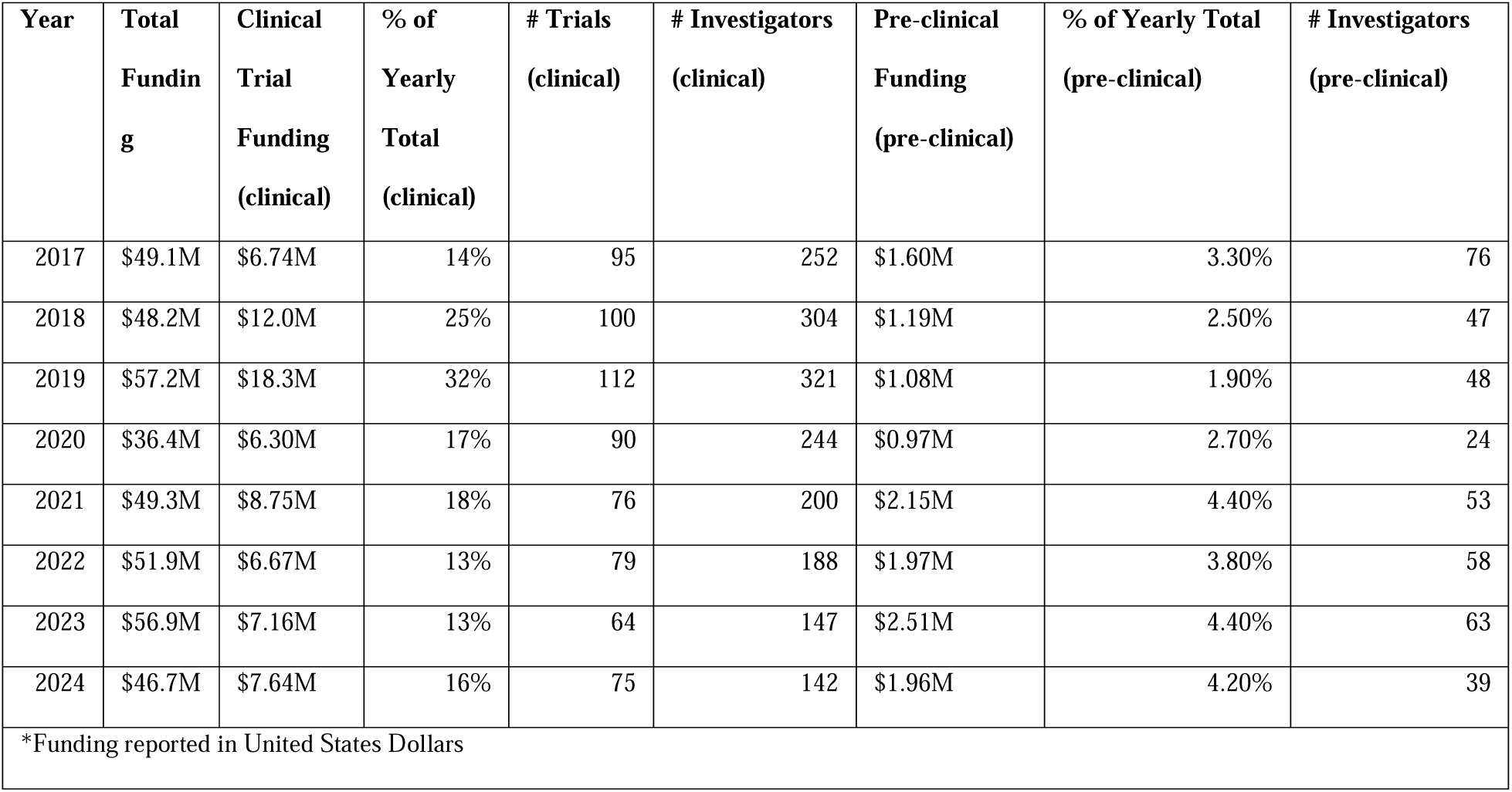
Designation of payments to clinical trials and pre-clinical research.

### Companies supplying research

From 2017-2024, 114 companies annually supported research to orthopaedic surgeons, ranging between 90 (2024) and 114 (2018–2019). Among the top 15 highest funding companies in 2024, ModernaTX was the top funder with approximately $5.3 million in ISRF, most of which ($4.76 million) was categorized as *unspecified*. Most companies funded device-oriented research, such as Arthrex, which was the second largest funder, with $4.0 million split between *device*-related ($1.49 million), *biological* ($198K), and *unspecified* ($2.33 million) support. The next six highest ISRF sponsors were almost exclusively *device* focused. However, several companies specializing in rare diseases, gene therapies, or personalized and regenerative medicine also composed top 15, including, Vertex, Ultragenyx, Medacta, and Miach.

From 2017 to 2024, device-oriented research consistently represented a major direction of ISRF, from a low of 43.5% in 2017 to a peak of 60.2% in 2020. Unspecified funding, which lacks a clear designation as drug, device, biological, or medical supply, remained substantial throughout the period, comprising 30–42% of annual totals. Biologics accounted for a variable portion of ISRF, from as high as 11.9% in 2021 to lows of 2–4%. Drug-related research peaked in 2019 (24.5%) but declined sharply thereafter, falling to just 1.4% in 2024, while medical supply funding was typically <1% (Figure 2B).

**Figure 2.**
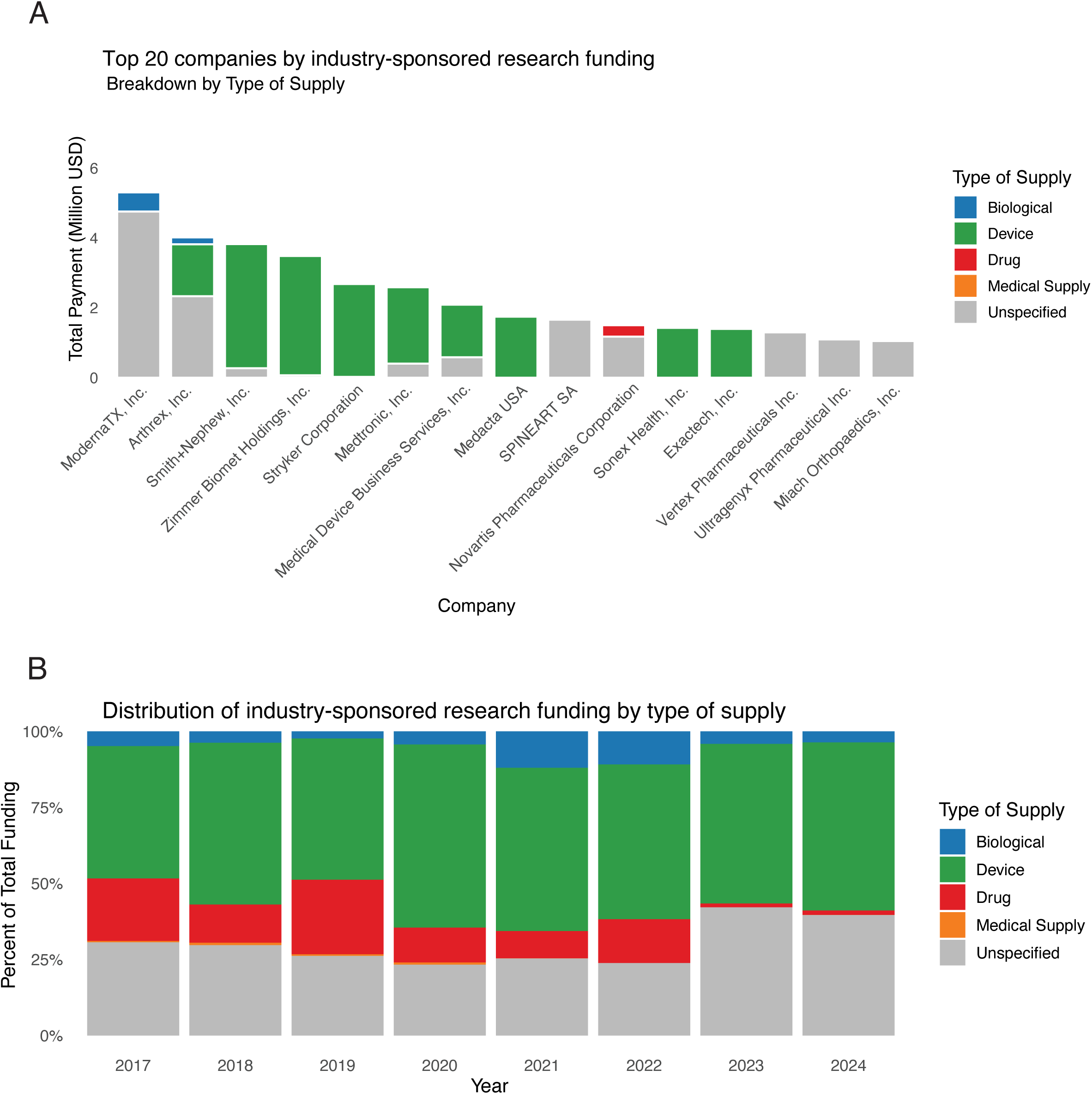
Designated Research Purpose of Research Funding. (A) The top 20 companies funding research to orthopaedic surgeons in 2024, highlighting their designed purpose which is terms the ‘Type of Supply’. (B) Each year, all industry related research funding was broken down by what percent of the total funding was going to each research category type of supply.

### Compensation for research

We next compared how orthopaedic surgeons who did and did not receive ISRF may also have been compensated by general payments. General payments to non-research-funded surgeons amounted to $167.5 million, whereas those with ISRF received $54.9 million. When assessed at the individual level in 2024, orthopaedic surgeons who received industry-sponsored research funding (ISRF) had substantially higher general (non-research) payments, averaging $69,441.35 with an interquartile range (IQR) of $219–$26,771. In contrast, the 20,032 orthopaedic surgeons without disclosed ISRF received an average general payment of $8,363.41 (IQR: $85–$1,041). Notably, the majority of orthopaedic surgeons in both groups received far less than the mean, with median payments of $1,714.83 among those with ISRF and $263.82 among those without ISRF (Figure 3). Using Cohen’s d effect size, this showed that those with ISRF receive a median general payments that are about 0.544 standard deviations higher than those who do not.

**Figure 3.**
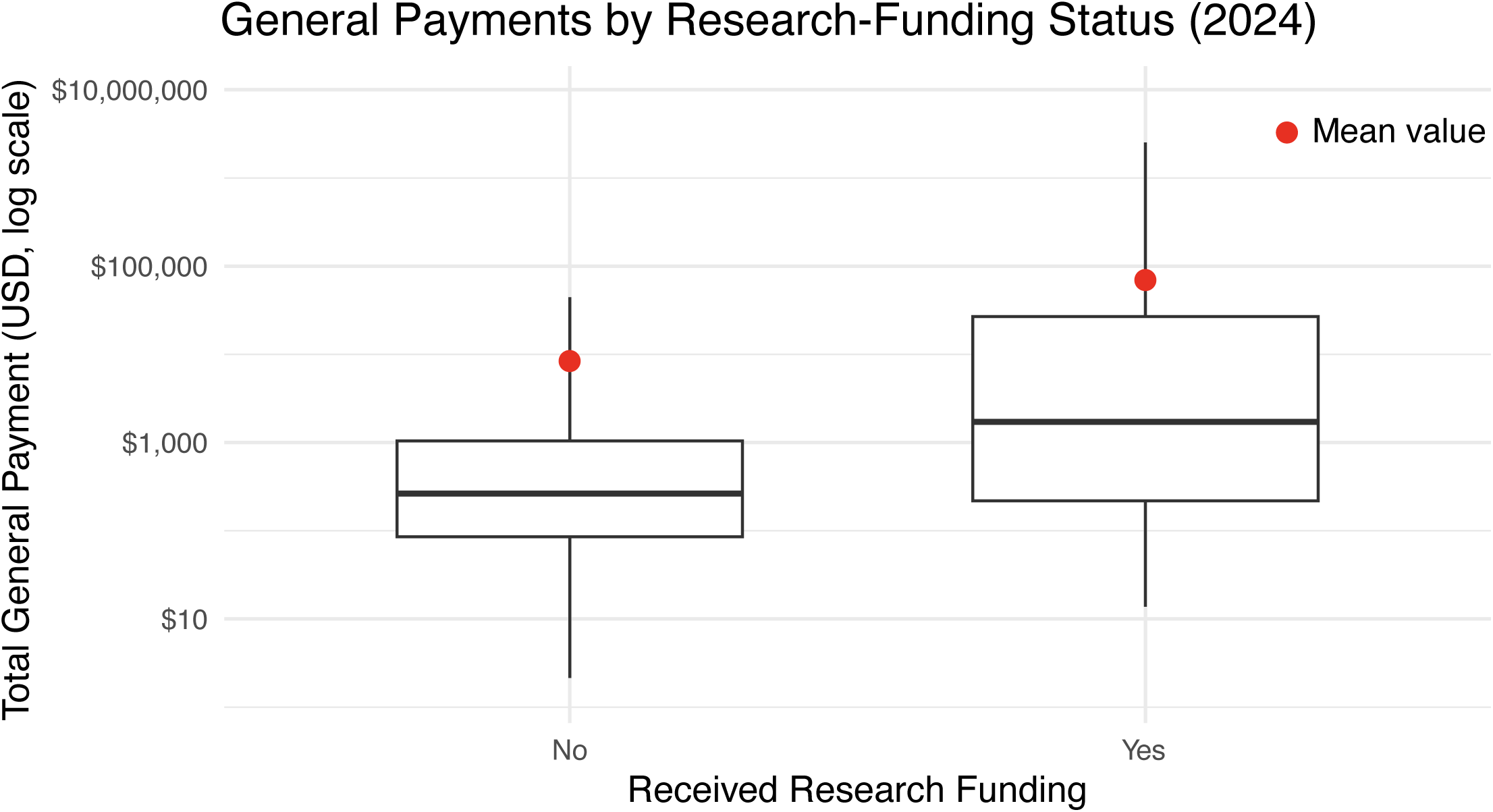
General payments to orthopaedic surgeons. In 2024, this histogram highlighting the inter-quartile range with the solid line as the median payment for those who did not ‘No’, and did ‘Yes’ receive research funding from an industry source. The y-axis is log(10) scaled in US Dollars. Center line inside the box is the median. Box edges are the 25th percentile (Q1) and 75th percentile (Q3). Whiskers extend to 1.5 × IQR from the box where outliers beyond these values are not included. Red dot indicates the median value received.

## Discussion

As federal funding available for MSK research is limited (1), ISRF serves as an additional resource along with grants directly from institutional and departmental sources, grants from orthopaedic societies/associations, and not-for-profit organizations. When ISRF funding is awarded to practicing orthopaedic surgeons, disclosures are made available through the Open Payments database supported by the Center for Medicare & Medicaid Services. Analyzing funding disclosure in this database to orthopaedic surgeons from 2017 to 2024, there was a total of $395,687,593 allocated during this eight-year period, averaging $49,460,949 annually, with a median funding of $14,121 per principal investigator (PI); these results align with a study assessing similar metrics up to 2021 (9). While most surgeons did not have an associated subspeciality listed (61.1%), there was no subspeciality that received significantly more funding than others. Some subspecialities, such as orthopaedic oncology, were not listed as they may have been grouped under the “unspecified” category. This is important to note since orthopaedic oncologists have some of the highest success rates at receiving external grants compared to other orthopaedic subspecialities (14).

While the median ISRF of ∼$14,000 is substantial, this was highly variable between individuals and is considerably lower than typical NIH grant amounts (estimated at $480,162 per award). However, very few orthopaedic surgeons receive NIH funding, as highlighted by a study that showed that only 43 orthopaedic surgeons received NIH support in 2022, totaling $21.1 million (15). This signifies that ISRF in orthopaedics provides broader, albeit smaller-scale, funding opportunities to more investigators than federal sources. That said, the reported data did not include the selection process or success rates for acquiring ISRF, which can be a major barrier to acquiring any research funding (14).

Federal research grants, namely from the NIH, primarily support pre-clinical (basic and translational) research. Within orthopaedic departments at US medical schools, 78.7% of NIH funding went toward supporting pre-clinical research, totaling $104,710,841 in 2021 (7). In contrast, industry support for pre-clinical research accounts for about 5% of allocated funding, whereas 13–32% of industry funding is associated with registered clinical trials, typically totaling around $8 million annually. For comparison, about 12.6% ($13.2 million) of NIH grants to orthopaedic departments are for clinical research (7).

Of the companies supporting ISRF in orthopaedics, the focus was weighted toward devices; a pattern that may be driven by market incentives and the regulatory advantages of the Food and Drug Administration (FDA)’s 510(k) clearance pathway. Because 510(k) approval relies on “substantial equivalence” to existing devices, it avoids the costly clinical trials required for pre-market approval (PMA) of high-risk devices, drugs, or biologics. (16,17). Not surprisingly, 88% of new orthopaedic devices, but only 53 % of devices in other fields, enter the market via 510(k). Others have shown that 510(k) product development recuperation costs are economically more favorable over PMAs (18,19), and may be a reason that about half of ISRF is directed towards devices, but only 13–32% of ISRF funding each year is associated with a clinical trial.

While device-focused ISRF accounted for about half the investments, several biotechnology companies were among the topic 15 funders. This could indicate future trends in orthopaedic treatment approaches as there is an emergence in regenerative and cell-based therapies. FDA approval of skeletal stem cells (20) and orthobiologics (21) in clinical practice will likely require a PMA and costly clinical trials prior to approval (16). Future ISRF may increasingly mirror specialties that rely heavily on clinical trials (PMAs), such as oncology, where ISRF to physicians approached $897 million in 2021 (22); far exceeding the $49 million that orthopaedic surgeons received in the same year. Continued revision and more stringent 510(k) guidelines could accelerate this transition (23) and lead to more high-quality clinical trials in orthopaedics (16).

The relative scarcity of research dollars in orthopaedics could be linked to the small pool of surgeon-scientists able, or willing, to conduct research (15,24,25). Compensation models that are generally dependent of clinical revenue can further drive active orthopaedic surgeons away from research due to the associated opportunity cost of time dedicated to research rather than clinical activities (26). Interestingly, orthopaedic surgeons serving as PIs for ISRF also receive a moderate statistically significant increase in general payments, averaging $61,078 higher than those who did not. This finding suggests that surgeons involved in research are leveraged by industry for their knowledge and skill set in other areas, as general payments largely consist of payments made for consulting or royalties. Furthermore, a study found a positive correlation between research productivity and compensation as well as ISRF (5,27). Together, compensation models by individual providers and academic institutions need to additionally factor in the holistic value of research. While these ISRF helps fill funding gaps and can help bring new therapies to market, they also raise ethical concerns about publishing bias and conflicts of interest (28). It has also been observed that self-reported conflicts diverge from Open Payments disclosures (29). Together, this signifies room to improve the Open Payments reporting in an effort to maintain public trust, promote transparency, and ensure patients get equitable, cost-effective care.

### Limitations

The data collected was based on discrete values and categories in the Open Payments database that were often categorical in nature. As our results showed, select categories were chosen to give a high-level overview of trends in ISRF, such as research category (e.g *Device, Biologic, Unspecific, etc*.). Other categories and details were often supplied but out of focus for this study, such as product category, specific device investigated, and name of study or individual clinical trial identification number. These caveats both highlight the limitations in the disclosures provided by the Open Payments database but also offer additional opportunities for analysis that extend beyond the overview in ISRF trends presented in this study. Additionally, as the data presented included data as part of disclosures made by healthcare providers, non-providers ISRF was not included. Non-MDs play substantial roles in advancing orthopaedic research, as highlighted by a study that showed within Departments of Orthopaedic Surgery at medical schools, a majority of funding is awarded to non-MDs (7,14).

### Conclusion

Our findings reveal that while ISRF in orthopaedics is widespread, it remains modest compared to federal funding levels and disproportionately directed toward device-related projects. This imbalance underscores a critical need for diversification of funding streams and stronger institutional support for clinical and translational orthopaedic research. Efforts to foster training pathways that incentivize surgeon participation in research is also critical. Future efforts should prioritize improving transparency and detail in Open Payments reporting, as evidenced by the large sums in ISRF categorized as unspecified. Strategic alignment between industry, academia, and federal agencies could ensure that orthopaedic research funding more accurately reflects the substantial disease burden and rapidly evolving therapeutic landscape of musculoskeletal care.

## Data Availability

All data is publicly available.

https://openpaymentsdata.cms.gov/datasets

## Acknowledgements and Disclosures

JSK received support from NIH T90 T90DE021989. AS received support from NIH R01DE030716. ILM industry disclosures include Orthozon, Spinal Simplicity, Stryker, ATEC, Spinewave, and Cerapedics.

